# Creation of an Open-Access Lung Ultrasound Image Database For Deep Learning and Neural Network Applications

**DOI:** 10.1101/2025.05.09.25327337

**Authors:** Andre Kumar, Pawan Nandakishore, Alexandra June Gordon, Evan Baum, Jai Madhok, Youyou Duanmu, John Kugler

**Author notes:** **Corresponding Author:** Andre Kumar MD, Stanford University School of Medicine, 300 Pasteur Drive, Stanford, CA 95401.

## Abstract

**Background:** Lung ultrasound (LUS) offers advantages over traditional imaging for diagnosing pulmonary conditions, with superior accuracy compared to chest X-ray and similar performance to CT at lower cost. Despite these benefits, widespread adoption is limited by operator dependency, moderate interrater reliability, and training requirements. Deep learning (DL) could potentially address these challenges, but development of effective algorithms is hindered by the scarcity of comprehensive image repositories with proper metadata.

**Methods:** We created an open-source dataset of LUS images derived a multi-center study involving N=226 adult patients presenting with respiratory symptoms to emergency departments between March 2020 and April 2022. Images were acquired using a standardized scanning protocol (12-zone or modified 8-zone) with various point-of-care ultrasound devices. Three blinded researchers independently analyzed each image following consensus guidelines, with disagreements adjudicated to provide definitive interpretations. Videos were pre-processed to remove identifiers, and frames were extracted and resized to 128×128 pixels.

**Results:** The dataset contains 1,874 video clips comprising 303,977 frames. Half of the participants (50%) had COVID-19 pneumonia. Among all clips, 66% contained no abnormalities, 18% contained B-lines, 4.5% contained consolidations, 6.4% contained both B-lines and consolidations, and 5.2% had indeterminate findings. Pathological findings varied significantly by lung zone, with anterior zones more frequently normal and less likely to show consolidations compared to lateral and posterior zones.

**Discussion:** This dataset represents one of the largest annotated LUS repositories to date, including both COVID-19 and non-COVID-19 patients. The comprehensive metadata and expert interpretations enhance its utility for DL applications. Despite limitations including potential device-specific characteristics and COVID-19 predominance, this repository provides a valuable resource for developing AI tools to improve LUS acquisition and interpretation.

## Introduction

Lung ultrasound (LUS) offers significant advantages as a bedside tool to diagnose pulmonary conditions.^1,2^ It can be expediently performed at the point of care to enhance diagnostic accuracy and monitor disease progression.^1,2^ LUS has demonstrated superior diagnostic accuracy compared to chest X-ray for identifying pneumonia, pulmonary edema, pleural effusion, and pneumothorax.^1,3–5^ When compared to computed tomography (CT), LUS often demonstrates similar diagnostic performance for these conditions.^6–8^ LUS devices are more cost-effective than traditional imaging equipment such as X-ray or CT machines, making them particularly valuable in resource-limited settings or overwhelmed clinical environments, as observed during the COVID-19 pandemic.^9,10^ Despite these advantages, widespread LUS adoption faces several challenges, including operator dependency for image acquisition, moderate interrater reliability for interpreting pathological findings, and the need for standardized provider training.^11,12^

Deep learning (DL), a subfield of artificial intelligence, has the potential to address many obstacles hindering LUS adoption.^13^ DL utilizes machine learning to automatically characterize features from raw data.^13–15^ By analyzing large repositories of ultrasound images, DL can make predictions about new images.^15^ Potential applications of DL in ultrasound include aiding examiners in image acquisition, providing anatomical labeling of structures, assessing image quality, identifying pathological findings, and assisting with interpretation.^13–16^ Through these capabilities, DL may improve LUS image quality, automate analysis, and help healthcare professionals make more accurate and timely clinical decisions.^13–15^

Despite the potential benefits of combining LUS with DL applications, there is a scarcity of high-quality image repositories needed to develop effective DL algorithms.^17–19^ There is a critical need to create open-source image libraries that can be used for current and future applications of ultrasound. Many existing databases derive primarily from inpatients and contain few healthy controls.^17,18^ Furthermore, these repositories often lack important metadata such as patient characteristics, scan locations, expert interpretations, and assessments of image quality.^17^

In this manuscript, we describe the creation of an open-source dataset of LUS images derived from N=226 patients, comprising N=303,977 individual video frames. This dataset represents one of the largest LUS repositories to date and includes comprehensive patient information, expert interpretations of findings, and control cases. Our dataset includes additional metadata to further aid in applications of DL as applied toward LUS.

## Methods

### Dataset Derivation

All patient images were collected as part of a multi-center prospective cohort study between 3/2020 and 4/2022 (Clinicaltrials.gov Registration: NCT04384055).^10,12^ Patient demographics and clinical information were collected. This study received institutional review board approval, including for the creation of the de-identified image database (Protocol 74680).

### Scanning Protocol

All images were acquired utilizing a standardized scanning protocol that has been previously published.^20^ Physicians were instructed to use a 12-zone scanning protocol for pulmonary views. If a 12-zone protocol could not be performed due to the patient’s physical condition, then a modified 8-zone protocol capturing the anterior and lateral lung fields was performed.^20^ This study utilized several POCUS devices, including Butterfly IQ™, Vave™, Sonosite™, GE Venue™ and Echonous™, which represent the commercially-available point-of-care machines at our institutions. All devices used a phased array probe and were set to the “lung” preset.

### Interpretation

Physician interpretation of the images was performed using a previously developed consensus guideline for LUS images.^21^ Three researchers (AK, JK, YD), who were blinded to any patient information, independently analyzed each image and provided their interpretation on separate electronic forms. The researchers then compared their assessments. When disagreements occurred, the researchers attempted to reach consensus. If no consensus could be reached, the interpretation for that image was marked as “indeterminate.” Previous investigations have demonstrated moderate to substantial interrater reliability (IRR) for LUS across different experience levels and probe types.^22,23^ In a previous analysis, we found that our interpretation protocol had moderate-to-substantial IRR for the LUS findings included in this study.^12^

### Image pre-processing

Each video was pre-processed using a pre-trained Unet model to remove device-specific identifiers, patient identifiers, and depth scales. OpenCV in Python (version 3.12.0) was used to extract frames from each video clip.^24^ Images were resized to 128×128 pixels for inference.

### Variable Definitions

All patient diagnoses were determined through retrospective chart reviews of the primary treating physician’s discharge summary or emergency department note. COVID-19 infection testing was obtained via nasopharyngeal polymerase chain reaction (PCR) in all patients presenting to the emergency department.

### Analysis

Summary statistics for the dataset, including mean, standard deviation (SD), and frequencies, were calculated. Chi-Square analysis was performed to compare frequencies of findings by lung zones. All analyses were conducted using Python (version 3.12.0).

## Results

The open access LUS repository and associated metadata can be found at: https://github.com/kumarandre/OpenPOCUS

The dataset consists of N=1,874 video clips (N=303,997 frames) derived from N=226 patients (Table 1). Of these participants, 114 (50%) were diagnosed with COVID-19 pneumonia at the time of their scan. Additional diagnoses are presented in Table 1. The mean body mass index (BMI) was 28.4 (SD 6.8), and 90 patients (40%) were female.

**Table 1.** Patient Demographics and Scan Characteristics. Other diagnoses included diarrhea N=2, osteomyelitis N=2, delirium N=2, failure to thrive N=3, stroke N=1, overdose N=1, acute coronary syndrome N=1, chest trauma N=4, and pleural effusion N=2.

|  |  |
| --- | --- |
| Patients, N | 226 |
| Video Clips, N | 1874 |
| Devices Used (%) |  |
| Butterfly | 134 (60) |
| Echonous | 21 (9.3) |
| Sonosite | 1 (0.4) |
| Vave | 56 (25) |
| GE Venue | 14 (6.2) |
| BMI, Mean (SD) | 28.4 (6.8) |
| Female, % | 90 (40) |
| Diagnosis (%) |  |
| COVID | 114 (50) |
| Bacterial Pneumonia | 12 (5.3) |
| Viral Pneumonia (non-COVID) | 12 (5.3) |
| Obstructive Airway Disease | 21 (9.3) |
| Healthy Control | 36 (16) |
| Pulmonary Edema | 14 (6.2) |
| Other | 17 (7.5) |

Regarding scanning protocols, 70 patients (31%) had scans obtained using the 12-zone protocol, while 156 patients (69%) had scans using the 8-zone protocol. Among all scans, 1,233 clips (66%) contained no abnormal findings, 338 clips (18%) contained B-lines, 84 clips (4.5%) contained consolidations, 119 clips (6.4%) contained both B-lines and consolidations, and 97 clips (5.2%) had indeterminate findings.

The dataset includes additional metadata such as COVID-19 status, normal versus abnormal interpretation, and expert interpretation of each video clip for a given lung zone (Table 2).

**Table 2.** Scan Counts and Findings. Each lung zone is displayed with the number of scans and findings.

|  | TOTAL | Zone 1 | Zone 2 | Zone 3 | Zone 4 | Zone 5 | Zone 6 | Zone 7 | Zone 8 | Zone 9 | Zone 10 | Zone 11 | Zone 12 |
| --- | --- | --- | --- | --- | --- | --- | --- | --- | --- | --- | --- | --- | --- |
| Scans | 1871 | 219 | 198 | 191 | 197 | 70 | 70 | 219 | 195 | 186 | 189 | 69 | 68 |
| Normal | 1233 | 154 | 137 | 123 | 136 | 43 | 39 | 144 | 140 | 106 | 122 | 46 | 43 |
| B-lines | 338 | 42 | 35 | 31 | 24 | 15 | 12 | 50 | 30 | 40 | 31 | 17 | 11 |
| Consolidation | 84 | 4 | 9 | 12 | 14 | 4 | 10 | 5 | 0 | 10 | 7 | 2 | 7 |
| B-lines & Consolidation | 119 | 17 | 15 | 19 | 6 | 3 | 4 | 14 | 5 | 19 | 11 | 1 | 5 |
| Indeterminate | 97 | 2 | 2 | 6 | 17 | 5 | 5 | 6 | 20 | 11 | 18 | 3 | 2 |

When categorized by lung zone of origin (Table 2), there were notable differences in the distribution of pathological findings. Anterior lung zones were more frequently interpreted as normal compared to lateral zones (70.3% vs. 64.3%; Chi-square 6.3, p=0.012) or posterior zones (70.3% vs. 61.5%, Chi-square 7.42; p<0.01; Supplementary Material). Consolidations were less frequently present in anterior lung zones compared to lateral zones (2.0% vs. 5.8%; Chi-Square 14.7; p<0.01) and posterior zones (2.0% vs. 8.3%, Chi-Square 22.9; p<0.01; Supplementary Material). Indeterminate findings occurred at low frequencies across all lung zone clips (Table 2).

## Discussion

We describe the creation of a comprehensive LUS dataset derived from adult patients with annotated metadata for each lung clip. This dataset encompasses patients with various pulmonary diseases as well as healthy controls. With over 303,997 individual frames, this open-source repository is among the largest available and uniquely includes patient-specific information, expert interpretations, and control cases. Together, the dataset can be used to aid in future applications of AI as applied toward lung ultrasound and medical imaging.

Previous investigations have found moderate to substantial interrater reliability for lung ultrasound findings,^12,23,25^ which is an important consideration given the expert over-reads of this study. Within our group, we have observed similar interrater reliability for the lung ultrasound findings in this database.^12^ For this project, we implemented an adjudication process to provide a single interpretation for future DL applications. As DL and neural network integration becomes more prominent in point-of-care ultrasound, determining how these technological advances can aid providers in image acquisition and interpretation will be crucial.^13,15^ It will be equally important to compare the accuracy of these models with expert physicians given these interrater reliabilities among providers are variable.^12,23,25^

Although other open-source LUS databases have been described, our dataset represents the largest to date.^26^ Few previous datasets contain annotated images.^26^ The majority of frames from these datasets are derived from patients with COVID-19.^17–19^ The heterogeneous manifestations of SARS-CoV-2 on lung parenchyma (including B-lines and consolidations) make it an ideal model for studying and developing lung ultrasound tools.^10^ However, as the pandemic evolves and COVID-19-related imaging becomes less frequent, there remains a critical need for datasets derived from other primary pulmonary pathologies. In our dataset, approximately half of the participants did not have COVID-19 at the time of their scan, representing a more diverse population compared to previously described datasets.^17–19,26^

It is important to note that there may be subtle differences in LUS findings between COVID-19 and other conditions. Models trained primarily on COVID-19 may miss these distinctions, particularly if used for diagnostic purposes.^14,26,27^ For example, cardiogenic pulmonary edema (due to heart failure) and non-cardiogenic pulmonary edema (related to acute respiratory distress syndrome, ARDS) present differently on LUS.^28,29^ Since COVID-19 typically causes imaging findings consistent with non-cardiogenic pulmonary edema, overreliance on COVID-19-derived models may reduce reliability in future imaging applications.^28,29^

### Limitations

This study has several limitations. We utilized only three reviewers for interpretation of each video clip, which may reduce the accuracy of findings. Analysis was performed at the clip level rather than the frame level, requiring caution when extrapolating metadata to individual frames. Approximately half of the scans were derived from COVID-19 patients, potentially limiting generalizability to specific disease models. The majority of scans (59%) were obtained using a single device (Butterfly IQ™). Despite pre-processing to remove device-specific markings, each ultrasound probe has unique characteristics (scan angle, frame rate, gray-scaling), potentially limiting model generalizability across devices.

### Conclusions

We present a large, open-source LUS database derived from diverse patients, including healthy controls. The methods used in creating this dataset can serve as a template for future datasets. Importantly, imaging datasets should be built on well-defined patient populations, which in this study included adults presenting to the emergency department with respiratory symptoms. Images should be reviewed by experts to label findings, and diagnoses should be assigned whenever possible. With the addition of similar LUS databases, it is possible to develop image interpretation tools powered by DL to assist in the acquisition and interpretation of point-of-care lung ultrasound.

## Data Availability

All data produced are available online at: https://github.com/kumarandre/OpenPOCUS

## Notes

### Competing Interest Statement

Andre Kumar reports consulting fees from Vave Health during the data collection period.

### Funding Statement

This study did not receive any funding

### Author Declarations

The Stanford University Institutional Review Board approved this study (Protocol 74680)

## References

1. Chiu L, Jairam MP, Chow R, et al. Meta-Analysis of Point-of-Care Lung Ultrasonography Versus Chest Radiography in Adults With Symptoms of Acute Decompensated Heart Failure. Am J Cardiol 2022;174:89–95.

2. Buhumaid RE, St-Cyr Bourque J, Shokoohi H, Ma IWY, Longacre M, Liteplo AS. Integrating point-of-care ultrasound in the ED evaluation of patients presenting with chest pain and shortness of breath. Am J Emerg Med 2019;37(2):298–303.

3. Pagano A, Numis FG, Visone G, et al. Lung ultrasound for diagnosis of pneumonia in emergency department. Intern Emerg Med 2015;10(7):851–4.

4. Perrone T, Maggi A, Sgarlata C, et al. Lung ultrasound in internal medicine: A bedside help to increase accuracy in the diagnosis of dyspnea. Eur J Intern Med 2017;46:61–5.

5. Dahmarde H, Parooie F, Salarzaei M. Accuracy of Ultrasound in Diagnosis of Pneumothorax: A Comparison between Neonates and Adults-A Systematic Review and Meta-Analysis. Can Respir J 2019;2019:5271982.

6. Nazerian P, Volpicelli G, Vanni S, et al. Accuracy of lung ultrasound for the diagnosis of consolidations when compared to chest computed tomography. Am J Emerg Med 2015;33(5):620–5.

7. Rocco M, Carbone I, Morelli A, et al. Diagnostic accuracy of bedside ultrasonography in the ICU: feasibility of detecting pulmonary effusion and lung contusion in patients on respiratory support after severe blunt thoracic trauma. Acta Anaesthesiol Scand 2008;52(6):776–84.

8. Chiumello D, Umbrello M, Sferrazza Papa GF, et al. Global and Regional Diagnostic Accuracy of Lung Ultrasound Compared to CT in Patients With Acute Respiratory Distress Syndrome. Crit Care Med 2019;47(11):1599–606.

9. Stanley A, Wajanga BMK, Jaka H, et al. The Impact of Systematic Point-of-Care Ultrasound on Management of Patients in a Resource-Limited Setting. Am J Trop Med Hyg 2017;96(2):488–92.

10. Kumar A, Weng Y, Duanmu Y, et al. Lung Ultrasound Findings in Patients Hospitalized With COVID-19. J Ultrasound Med [Internet] 2021;Available from: 10.1002/jum.15683

11. Kumar, A., Jensen, T., Kugler, J. Evaluation of trainee competency with point-of-care ultrasonography (POCUS): a conceptual framework and review of existing assessments. J Gen Intern Med 2019;[Epub ahead of print].

12. Kumar A, Weng Y, Graglia S, et al. Interobserver agreement of lung ultrasound findings of COVID-19. J Ultrasound Med [Internet] 2021; Available from: 10.1002/jum.15620

13. Shokoohi H, LeSaux MA, Roohani YH, Liteplo A, Huang C, Blaivas M. Enhanced Point-of-Care Ultrasound Applications by Integrating Automated Feature-Learning Systems Using Deep Learning. J Ultrasound Med 2019;38(7):1887–97.

14. Liu S, Wang Y, Yang X, et al. Deep Learning in Medical Ultrasound Analysis: A Review. Proc Est Acad Sci Eng 2019;5(2):261–75.

15. Baum E, Tandel MD, Ren C, et al. Acquisition of cardiac point-of-care ultrasound images with deep learning. CHEST Pulmonary 2023;(100023):100023.

16. Nti B, Lehmann AS, Haddad A, Kennedy SK, Russell FM. Artificial Intelligence-Augmented Pediatric Lung POCUS: A Pilot Study of Novice Learners. J Ultrasound Med 2022;41(12):2965–72.

17. Born J, Wiedemann N, Cossio M, et al. Accelerating detection of lung pathologies with explainable ultrasound image analysis. Appl Sci (Basel) 2021;11(2):672.

18. Etter L, Betke M, Camelo IY, et al. Curated and annotated dataset of lung US images in Zambian children with clinical pneumonia. Radiol Artif Intell 2024;6(2):e230147.

19. Ebadi A, Xi P, MacLean A, et al. COVIDx-US: An open-access benchmark dataset of ultrasound imaging data for AI-driven COVID-19 analytics. Front Biosci (Landmark Ed) 2022;27(7):198.

20. Doerschug KC, Schmidt GA. Intensive care ultrasound: III. Lung and pleural ultrasound for the intensivist. Ann Am Thorac Soc 2013;10(6):708–12.

21. Kumar A, Weng I, Graglia S, et al. Point-of-Care Ultrasound Predicts Clinical Outcomes in Patients With COVID-19. J Ultrasound Med [Internet] 2021; Available from: 10.1002/jum.15818

22. Gomond-Le Goff C, Vivalda L, Foligno S, Loi B, Yousef N, De Luca D. Effect of Different Probes and Expertise on the Interpretation Reliability of Point-of-Care Lung Ultrasound. Chest 2020;157(4):924–31.

23. Vieira JR, Castro MR de, Guimarães T de P, et al. Evaluation of pulmonary B lines by different intensive care physicians using bedside ultrasonography: a reliability study. Rev Bras Ter Intensiva 2019;31(3):354–60.

24. Bradski G. The opencv library. Dr Dobb’s Journal: Software Tools for the Professional Programmer 2000;25(11):120–3.

25. Gullett J, Donnelly JP, Sinert R, et al. Interobserver agreement in the evaluation of B-lines using bedside ultrasound. J Crit Care 2015;30(6):1395–9.

26. Wang J, Yang X, Zhou B, et al. Review of machine learning in lung ultrasound in COVID-19 pandemic. J Imaging 2022;8(3):65.

27. Cheema BS, Walter J, Narang A, Thomas JD. Artificial Intelligence-Enabled POCUS in the COVID-19 ICU: A New Spin on Cardiac Ultrasound. JACC Case Rep 2021;3(2):258–63.

28. Sekiguchi H, Schenck LA, Horie R, et al. Critical care ultrasonography differentiates ARDS, pulmonary edema, and other causes in the early course of acute hypoxemic respiratory failure. Chest 2015;148(4):912–8.

29. Heldeweg MLA, Smit MR, Kramer-Elliott SR, et al. Lung Ultrasound Signs to Diagnose and Discriminate Interstitial Syndromes in ICU Patients: A Diagnostic Accuracy Study in Two Cohorts. Crit Care Med 2022;50(11):1607–17.

