## Supplementary material for "Creation of an Open-Access Lung Ultrasound Image Database For Deep Learning and Neural Network Applications": Supplementary Material.docx

| Pathology | Lung Areas | Number | p-value |
| --- | --- | --- | --- |
| Normal | Anterior | 575 | 0.692 |
|  | Lateral | 505 | 0.662 |
|  | Posterior | 171 | 0.617 |
|  | Left | 601 | 0.649 |
|  | Right | 632 | 0.669 |

| Pathology | Lung Areas | Number | p-value |
| --- | --- | --- | --- |
| B-lines | Anterior | 157 | 0.189 |
|  | Lateral | 126 | 0.165 |
|  | Posterior | 55 | 0.199 |
|  | Left | 179 | 0.193 |
|  | Right | 159 | 0.168 |

| Pathology | Lung Areas | Number | p-value |
| --- | --- | --- | --- |
| Consolidation | Anterior | 18 | 0.022 |
|  | Lateral | 43 | 0.056 |
|  | Posterior | 23 | 0.083 |
|  | Left | 31 | 0.033 |
|  | Right | 53 | 0.056 |

| Pathology | Lung Areas | Number | p-value |
| --- | --- | --- | --- |
| Both | Anterior | 51 | 0.061 |
|  | Lateral | 55 | 0.072 |
|  | Posterior | 13 | 0.047 |
|  | Left | 55 | 0.059 |
|  | Right | 64 | 0.068 |

| Pathology | Lung Areas | Number | p-value |
| --- | --- | --- | --- |
| Indeterminate | Anterior | 30 | 0.036 |
|  | Lateral | 52 | 0.068 |
|  | Posterior | 15 | 0.054 |
|  | Left | 60 | 0.065 |
|  | Right | 37 | 0.039 |

Supplementary Tables: Pathology by Lung Areas. Chi-Square analysis was used for calculating the different proportions of pathological findings by lung zones.
